# Archi-Prevaleat project. A National cohort of color-Doppler ultrasonography of the epi-aortic vessels in Patients Living with HIV

**DOI:** 10.1101/2022.08.02.22278263

**Authors:** P. Maggi, E.D. Ricci, C. Muccini, L. Galli, B.M. Celesia, S. Ferrara, Y. Salameh, R. Basile, G. Di Filippo, F. Taccari, A. Tartaglia, A. Castagna

## Abstract

**Objectives:** To evaluate the prevalence of carotid intima-media thickness and plaques in a cohort of persons living with HIV, the role of cardiovascular risk factors, the impact of the antiretroviral regimens, and the difference between naïve and experienced patients in the onset of carotid lesions.

**Methods:** This project was initiated in 2019 and involves eight Italian Centers. Carotid changes were detected using a power color-Doppler ultrasonography with 7.5 MHz probes. The following parameters are evaluated: intima-media thickness of both the right and left common and internal carotids: Data regarding risk factors for CVD, HIV viral load, CD4+ cell counts, serum lipids, glycaemia, and body mass index. The associations between pathological findings and potential risk factors were evaluated by logistical regression, with odds ratios (OR) and 95% confidence intervals (95% CI).

**Results:** Among 1147 evaluated patients, aged 52 years on average, 347 (30.2%) had pathological findings (15.8% plaques and 14.5% IMT). Besides usual risk factors, such as older age, male sex, and dyslipidemia, CD4+ cell nadir <200 cells/mL (OR 1.51, 95% CI 1.14-1.99) and current use of raltegravir (OR 1.54, 95% CI 1.01-2.36) were associated with higher prevalence of pathological findings.

**Conclusions:** Our data show that the overall percentage of carotid impairments nowadays remains high. Color-Doppler ultrasonography could play a pivotal role in identifying and quantifying atherosclerotic lesions among persons living with HIV, even at a very premature stage, and should be included in the algorithms of comorbidity management of these patients.

## Introduction

The introduction of effective antiretroviral (ARV) regimens had a deep impact on the natural history of HIV infection, leading to a dramatic decrease in its mortality rate and a considerable increase in the life expectancy of Persons Living with HIV (PLWH). Nevertheless, these patients still appear to be at higher risk of developing several co-morbidities, such as cardiovascular disease (CVD) [1, 2]. Measurement of the carotid intima-media thickness (IMT) and detection of plaques with color-Doppler ultrasonography is a non-invasive, sensitive, and highly reproducible technique aimed not only at assessing vascular anatomy and function, but also at identifying and quantifying atherosclerotic lesions, even at a very premature stage. It is a well-validated research tool and is widely used in clinical practice [3]. This technique allows the measurement of a variety of parameters including the IMT, arterial diameter, presence of plaques, as well as blood flow and velocity.

PREVALEAT (PREmature VAscular LEsions and Antiretroviral Therapy) is an ongoing multicenter, longitudinal cohort involving several Italian centers since 1998, aimed at evaluating the cardiovascular (CV) risk in PLWH, using color-Doppler ultrasonography. The cohort resulted in several studies being done in this field, throughout the years, [4–7]. Considering that the use of this technique is, at present, widely diffused among the Italian HIV outpatient facilities, we generated a National cohort of color-Doppler ultrasonography (Archi-Prevaleat) to better evaluate the characteristics of vascular lesions in PLWH, based on a large amount of data.

The aim of the present study is to evaluate the prevalence of carotid IMT and plaques in this cohort of PLWH, the role of classic CV risk factors, and of the risk factors related to HIV infection, as well as the impact of the different antiretroviral regimens, and finally, the difference between naïve and experienced patients in the onset of these carotid lesions.

## Patients and methods

This ongoing project was initiated in 2019 and involves, currently, eight Italian Infectious Diseases Centers in which the ultrasonographic examination is performed by competent physicians. These physicians were specifically trained on the technique during a Continuing Medical Education training program organized by the former coordinating Center (Azienda Ospedaliera Ospedale Policlinico Consorziale Bari – Italy) until 2021 and, at present, by the current coordinating Center (Università della Campania, Luigi Vanvitelli). Comparison and standardization of the technique is performed using images and filmed reports during annual follow up meetings. Written informed consent was obtained from each patient. The study protocol conforms to the ethical guidelines of the 1975 Declaration of Helsinki. The central Ethics Committee (Comitato Etico Indipendente Locale – Azienda Ospedaliera Ospedale Policlinico Consorziale Bari – Italy) gave ethical approval for this work, within the PREVALEAT cohort study protocol, active since 1998 (last amendment 2019).

The cohort’s data was registered on an online platform (http://www.archiprevaleat.com/). The data collected concerns patients when they presented to the examination for the first time and routinely at all the subsequent follow up examinations

The following parameters are evaluated:

1. IMT of both the right and left common and internal carotids: ultrasonography of the epi-aortic vessels is performed using a power color-Doppler instrument with 7.5 MHz probes. The characteristics of the intima together with the pulsation index, the resistance index, the minimal speed, the peak speed, and the mean speed will be evaluated. A minimum of three measurements is requested: on the common carotid artery, 1 cm before the carotid bifurcation and at the carotid bifurcation; and also on the internal carotid, 1 cm after the carotid bifurcation and 2 cm after the carotid bifurcation. An IMT >1.0 mm is to be considered pathological. Atherosclerotic plaques, if present, are described. A carotid was classified as being affected by plaques if there was a localized thickening >1.2 mm that did not uniformly involve the whole left or right common carotid bifurcation with or without flow disturbance (8, 9). All relevant images are photographed and properly archived.
2. Data regarding risk factors for CVD (family history, smoke, active drug addiction, alcohol consumption) are to be collected at baseline, and re-evaluated every 12 months.
3. HIV viral load, CD4+ cell counts, serum total cholesterol, low density lipoprotein cholesterol (LDL-c), high density lipoprotein cholesterol (HDL-c), glycaemia, triglycerides, and body mass index (BMI) are to be recorded at every annual control.

### Statistical methods

Categorical and ordinal variables were described as frequency (%) and were compared between groups using the heterogeneity χ2 test (or the Mantel-Hanszel χ2 test as appropriate). Numerical variables were described as mean, and standard deviation (SD) if normally distributed, or median, and interquartile range (IQR) if not normally distributed. Comparisons were performed using the analysis of variance, the Mann-Whitney test (or the Kruskal-Wallis test as appropriate), respectively. Odds ratios (ORs) and the corresponding 95% confidence intervals (CI) were used to evaluate the association between IMT and patients’ characteristics and clinical variables. Two models for multivariate analyses were opted to select for potential confounders. All variables, that were significantly associated with pathological results at the univariate analysis, will be controlled in a model including sex and age. Variables still significantly associated with IMT in this analysis were included in the final model. To avoid losing observations, classes were created for missing values of selected variables.

Data analysis was conducted with SAS for Windows 9.4 (SAS Institute Inc., Cary, NC, USA).

## Results

1147 patients who underwent color-Doppler ultrasonography in the participating Centers (82% males, mean age 52.1, SD 10.0 years) were enrolled. Prevalence of IMT and plaques at the left carotid were 12.1% (n=139) and 12.5% (n=143) respectively, and 11.2% (n=129) and 10.1% (n=116) at the right carotid, respectively (Fig. 1). The overall percentage of patients with carotid impairments (IMT or plaques either in the right or left carotid) was 30.2% (n=347) while the remaining 69.8% (n=800) were normal. Among pathologic patients, 15.8% (n=181) had plaques, while 14.5% (n=166) had only IMT. Demographic and metabolic data, data regarding HIV infection and cardio-cerebral-vascular comorbidities are summarized in Table 1.

**Figure 1.**
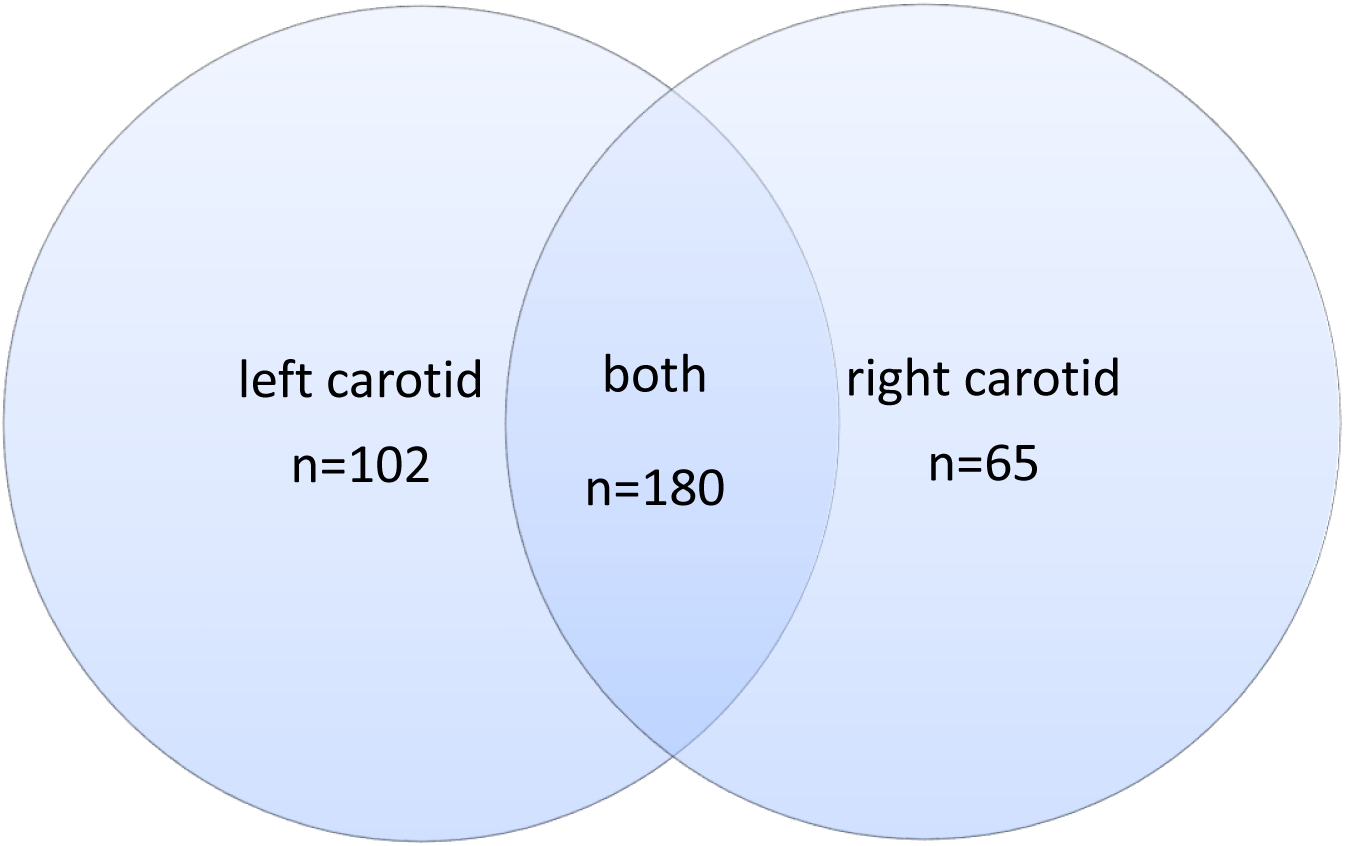
Pathological findings at left and right carotids.

**Table 1.**
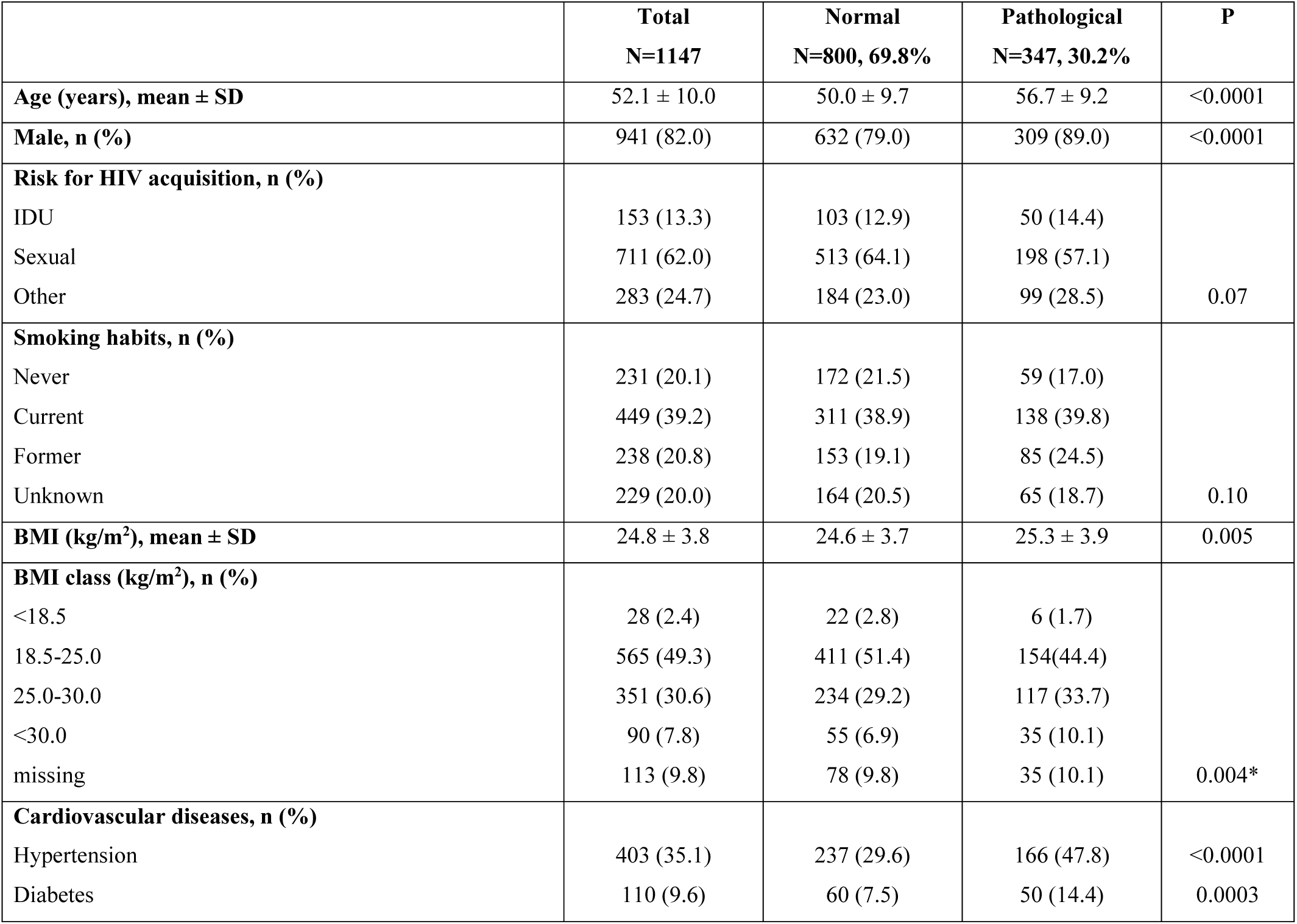
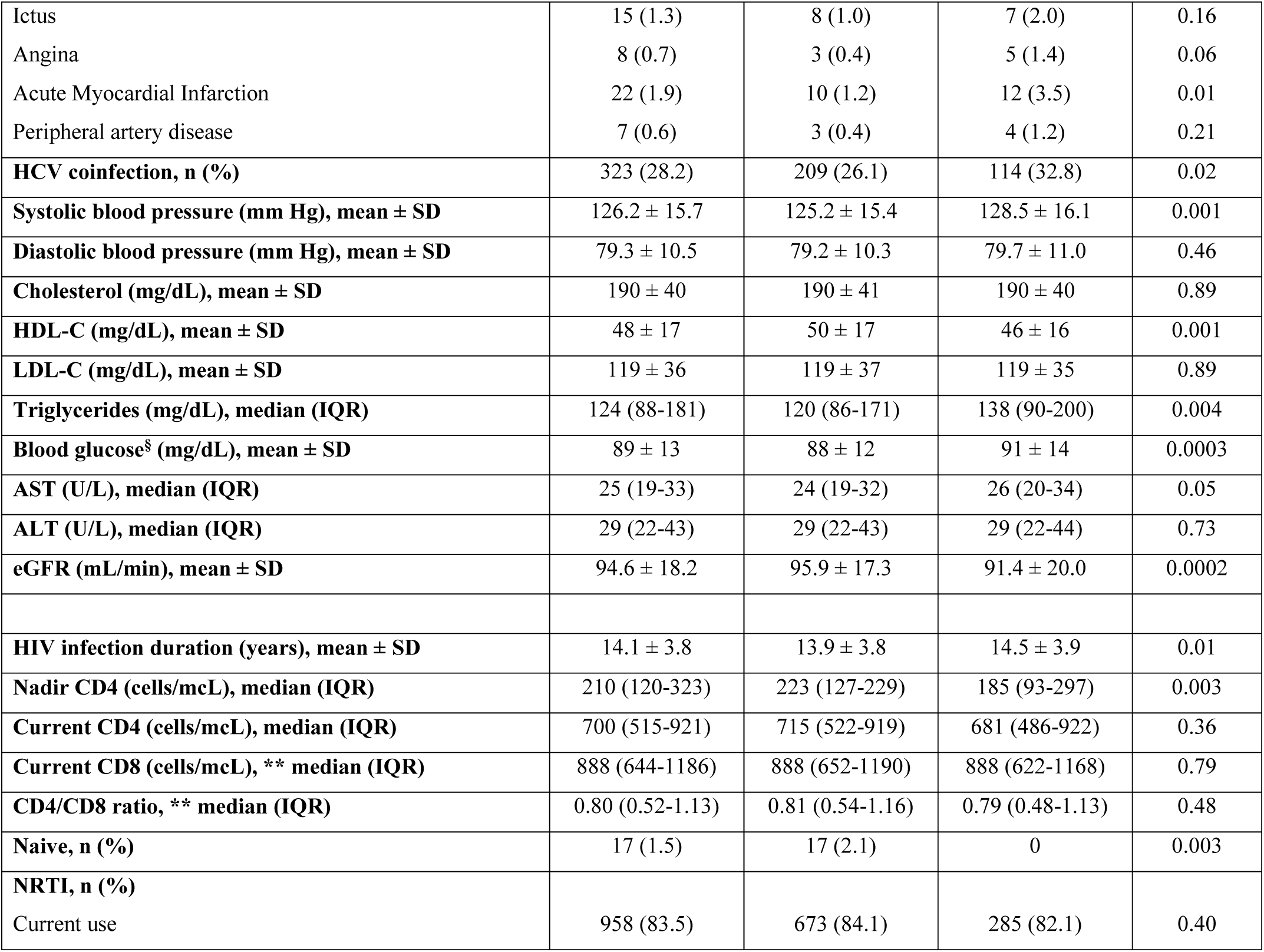
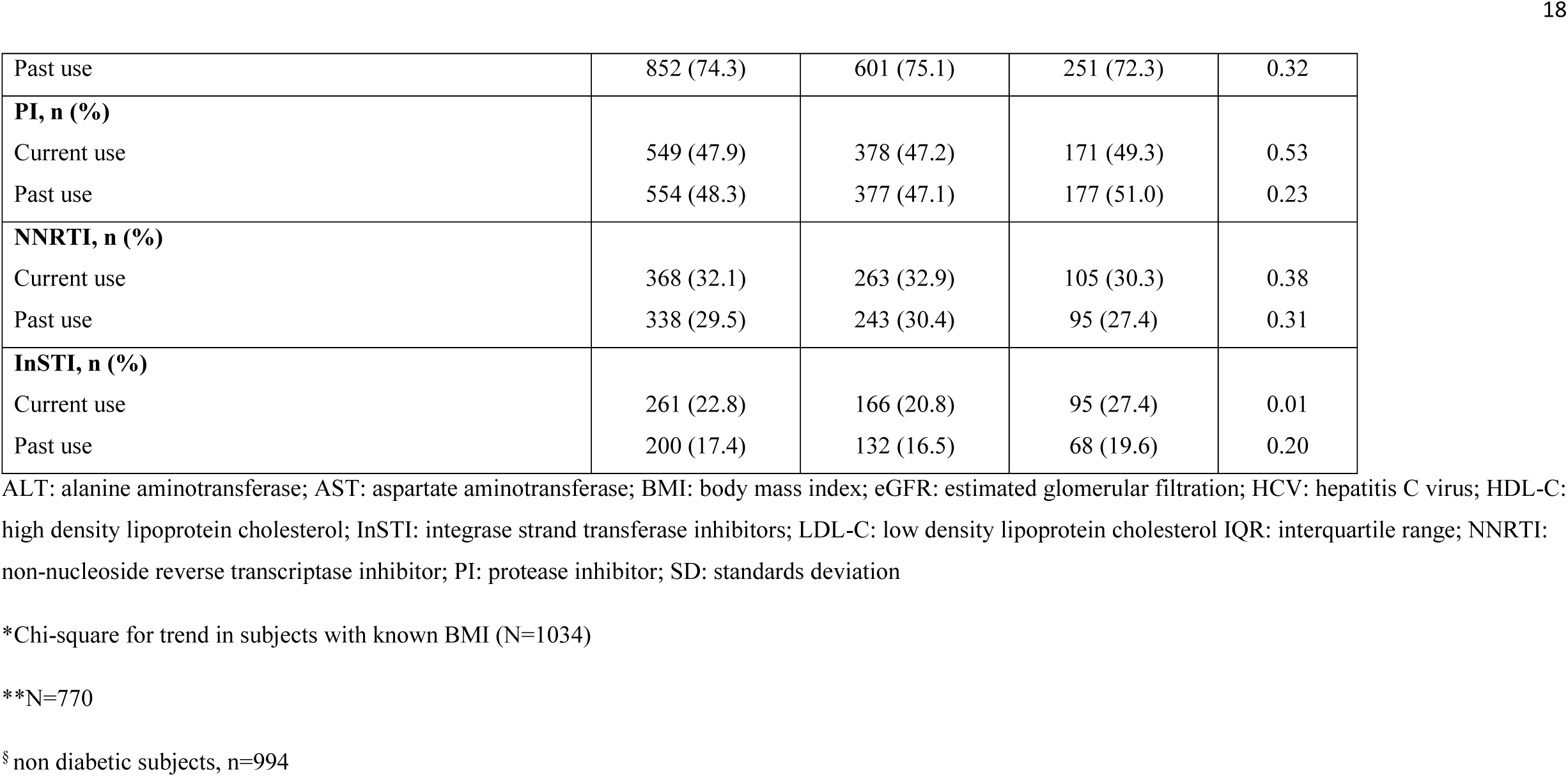
Main characteristics of 1147 HIV-positive patients, according to the presence of pathological results (IMT> 1.0 mm).

We observed that, regarding demographics, carotid impairments were significantly related to age (p<0.0001), male sex (p<0.0001), and BMI (p=0.005). Regarding comorbidities, individuals with pathological findings had: i) more frequent prevalence of hypertension (p<0.0001), diabetes (p=0.0003), previous myocardial infarction (p=0.01) and HCV coinfection (p=0.02); ii) higher values of systolic blood pressure (p=0.001), triglycerides (p<0.0001), blood glucose, regardless of a diagnosis of diabetes (p=0.0003); iii) lower values of HDL-cholesterol (p=0.001), and eGFR (p<0.0001) than individuals with normal findings. Considering variables related to HIV infection, there was a significant relationship between the presence of IMT and/or plaques and a longer HIV infection duration (p=0.01), a lower CD4+ cell nadir (p=0.003) and a current use of integrase inhibitors (p=0.003). Exploring details of current antiretroviral treatments, we found that the current use of raltegravir is significantly associated with pathological findings (p=0.0001), as shown in Table 2. On the contrary, a naïve status was related with normal findings (p=0.01), although this result emerged from a limited sample of 17 patients.

**Table 2.** Details of most frequently used ARV treatment in 1147 HIV-positive patients, according to the presence of plaques.

|  | **Total**<br>**N=1147** | **Normal**<br>**N=800, 69.8%** | **Pathological**<br>**N=347, 30.2%** | **P** |
| --- | --- | --- | --- | --- |
| **Current NRTI, n (%)** |  |  |  |  |
| Tenofovir | 539 (47.0) | 380 (47.5) | 159 (45.8) | 0.60 |
| Abacavir | 280 (24.4) | 198 (24.7) | 82 (23.6) | 0.68 |
| Lamivudine | 420 (36.6) | 294 (36.8) | 126 (36.3) | 0.89 |
| Emtricitabine | 526 (45.9) | 375 (46.9) | 151 (43.5) | 0.29 |
| **Current PI, n (%)** |  |  |  |  |
| Atazanavir | 252 (22.0) | 175 (21.9) | 77 (22.2) | 0.91 |
| Darunavir | 248 (21.6) | 169 (21.1) | 79 (22.8) | 0.54 |
| **Current NNRTI, n (%)** |  |  |  |  |
| Efavirenz | 144 (12.6) | 103 (12.9) | 41 (11.8) | 0.62 |
| **Current InSTI, n (%)** |  |  |  |  |
| Raltegravir | 119 (10.4) | 65 (8.1) | 54 (15.6) | 0.0001 |
| Dolutegravir | 102 (8.9) | 72 (9.0) | 30 (8.6) | 0.85 |
| Elvitegravir | 40 (3.5) | 29 (3.6) | 11 (3.2) | 0.70 |

The crude ORs, sex- and age-adjusted ORs (aOR) and the complete model aOR (with corresponding 95% CI) are reported in Table 3. The associations were shown to be statistically significant for age (by 1 year, OR 1.08 and aOR 1.08, respectively), female sex (OR 0.46 and aOR 0.52), BMI (by 1 kg/m2, OR 1.05 and aOR 1.05) and >30.0 (OR 1.70 and aOR 1.60), hypertension (OR 2.18 and aOR 1.45), HCV coinfection (OR 1.38 and aOR 1.44), elevated triglycerides (OR 1.64 and aOR 1.45), low HDL-c (OR 1.46 and aOR 1.64), INSTI use (OR 1.44 and aOR 1.38) and raltegravir use (OR 2.08 and aOR 1.74), respectively in the crude and sex- and age-adjusted analyses.

**Table 3.** Odds ratios (OR) and 95% confidence interval (95% CI), crude and sex- and age-adjusted.

|  | Crude |  |  | Age- and sex-adjusted |  |  | Complete model** |  |  |
| --- | --- | --- | --- | --- | --- | --- | --- | --- | --- |
|  | OR | 95% CI |  | aOR | 95% CI |  | aOR | 95% CI |  |
| **Age (by 1 year)** | **1.08** | **1.06** | **1.09** | **1.08** | **1.06** | **1.09** | **1.07** | **1.06** | **1.09** |
| **Female (ref. Male)** | **0.46** | **0.32** | **0.68** | **0.52** | **0.35** | **0.77** | **0.54** | **0.36** | **0.81** |
| **Smoking habits (ref. Never)** |  |  |  |  |  |  |  |  |  |
| Current | 1.29 | 0.90 | 1.85 | 1.35 | 0.92 | 1.97 |  |  |  |
| Former | **1.62** | **1.09** | **2.41** | 1.31 | 0.86 | 2.06 |  |  |  |
| Unknown | 1.16 | 0.76 | 1.74 | 1.41 | 0.91 | 2.20 |  |  |  |
| **BMI (by 1 kg/m<sup>2</sup>)** | **1.05** | **1.01** | **1.09** | **1.05** | **1.01** | **1.09** |  |  |  |
| **BMI class (ref. 18.6-25.0 kg/m<sup>2</sup>)** |  |  |  |  |  |  |  |  |  |
| <18.5 | 0.73 | 0.29 | 1.83 | 0.71 | 0.26 | 1.89 | 0.77 | 0.29 | 2.07 |
| >25.0-30.00 | **1.33\*** | **1.00** | **1.78** | 1.32 | 0.97 | 1.80 | 1.28 | 0.93 | 1.75 |
| >30.00 | **1.70** | **1.07** | **1.70** | **1.60** | **1.04** | **2.67** | 1.50 | 0.90 | 2.49 |
| **Hypertension** | **2.18** | **1.68** | **2.82** | **1.45** | **1.09** | **1.92** | 1.28 | 0.95 | 1.72 |
| **Diabetes** | **2.08** | **1.39** | **3.09** | 1.29 | 0.84 | 1.97 |  |  |  |
| **Angina** | 3.88 | 0.92 | 16.34 | 1.91 | 0.79 | 4.60 |  |  |  |
| **IMA** | **2.83** | **1.21** | **6.61** | 1.30 | 0.54 | 3.16 |  |  |  |
| **HCV coinfection (ref. N)** | **1.38** | **1.05** | **1.82** | **1.44** | **1.08** | **1.93** | 1.34 | 0.99 | 1.82 |
| **HDL-C (ref. ≥40 (M) ≥50 (F) mg/dL)** | **1.46** | **1.10** | **1.93** | **1.64** | **1.22** | **2.22** | **1.45** | **1.06** | **2.00** |
| **Triglycerides (ref. <150 mg/dL)** | **1.64** | **1.26** | **2.14** | **1.45** | **1.10** | **1.92** | 1.21 | 0.90 | 1.64 |
| **Blood glucose (ref. <100 mg/dL)** | **1.62** | **1.21** | **2.18** | 0.95 | 0.69 | 1.32 |  |  |  |
| **Systolic blood pressure (by 5 mm Hg)** | **1.07** | **1.02** | **1.11** | 1.01 | 0.97 | 1.03 |  |  |  |
| **eGFR (by 5 ml/min)** | **0.94** | **0.90** | **0.97** | 1.03 | 0.98 | 1.07 |  |  |  |
| **HIV infection duration (by 5 years)** | **1.26** | **1.06** | **1.50** | 1.01 | 0.82 | 1.23 |  |  |  |
| **Nadir CD4 (ref. $\geq 200$ cells/mm<sup>3</sup>)** | **1.48** | **1.14** | **1.90** | **1.58** | **1.21** | **2.07** | **1.51** | **1.14** | **1.99** |
| **Current INSTI use (ref. N)** | **1.44** | **1.08** | **1.93** | **1.38** | **1.02** | **1.89** | 1.28 | 0.93 | 1.77 |
| **Current raltegravir use (ref. N)** | **2.08** | **1.42** | **3.06** | **1.74** | **1.16** | **2.61** | **1.54** | **1.01** | **2.36** |

**Bold:** $p < 0.05$
**\* $p = 0.05$**
**\*\*** aOR: included all variables with $p < 0.05$ at the age- and sex-adjusted analysis.
Estimates for Raltegravir use and INSTI use were derived from separate models.

The previous smoking (OR 1.62), myocardial infarction (OR 2.83), diabetes (OR 2.08), blood glucose (OR 1.62), eGFR (OR 0.94), and HIV infection duration (OR 1.26) were only significant in the crude model.

Thus, the final model was made to include age, sex, BMI, hypertension, HCV coinfection, HDL, triglycerides, nadir CD4 and, in turn, INSTI use or raltegravir use. In this model, age, sex, HDL-C, nadir CD4 and raltegravir use, maintained their association with a pathological IMT (Table 3).

Dividing the pathological findings into two classes, IMT between >1.00 and 1.20 mm and presence of plaques, the analyses were rerun. At univariate, the associated variables were age, sex, BMI, hypertension, diabetes, angina, acute myocardial infarction, HDL-C, triglycerides, nadir CD4 <200, previous NNRTI use, current INSTI use, and current raltegravir use. After adjusting for age and sex, the BMI, hypertension, diabetes, lowered HDL-C, increased TGL, nadir CD4 <200, current INSTI use, and current RAL use were associated to presence of plaques. IMT between >1.00 and 1.20 mm was associated to HCV coinfection.

Thus, all these factors were included in the complete models, for risk of IMT and plaques, INSTI and RAL use were included in different models). Age and sex were significantly associated with pathological findings in both classes, whereas BMI, HDL-C, TGL and nadir CD4 were only significant in presence of plaques (Table 4).

**Table 4.** Adjusted odds ratios (OR) and 95% confidence interval (95% CI).

|  | IMT 1.01-1.20 |  |  | Plaques |  |  |
| --- | --- | --- | --- | --- | --- | --- |
|  | aOR | 95% CI |  | aOR | 95% CI |  |
| **Age (by year)** | **1.06** | **1.03** | **1.08** | **1.09** | **1.07** | **1.11** |
| **Female (ref. Male)** | **0.56** | **0.33** | **0.94** | **0.56** | **0.31** | **0.98** |
| **BMI (ref. 18.6-25.0 kg/m<sup>2</sup>)** |  |  |  |  |  |  |
| ≤18.5 | 0.48 | 0.10 | 2.14 | 1.13 | 0.34 | 3.73 |
| >25.0 | 1.06 | 0.72 | 1.56 | **1.65** | **1.12** | **2.45** |
| **Hypertension** | 1.24 | 0.85 | 1.82 | 1.32 | 0.90 | 1.95 |
| **Diabetes** | 0.73 | 0.39 | 1.38 | 1.27 | 0.74 | 2.18 |
| **HCV coinfection (ref. N)** | 1.41 | 0.97 | 2.06 | 1.26 | 0.84 | 1.89 |
| **HDL-C (ref. ≥40 (M) ≥50 (F) mg/dL)** | 1.21 | 0.80 | 1.82 | **1.77** | **1.17** | **2.68** |
| **Triglycerides (ref. <150 mg/dL)** | 0.92 | 0.62 | 1.36 | **1.66** | **1.12** | **2.46** |
| **Nadir CD4 (ref. ≥ 200 cells/mm<sup>3</sup>)** | 1.19 | 0.83 | 1.69 | **1.79** | **1.23** | **2.70** |
| **Past NNRTI (ref. N)** | 1.10 | 0.75 | 1.59 | 0.68 | 0.44 | 1.05 |
| **Current raltegravir use (ref. N)** | 1.61 | 0.95 | 2.74 | 1.55 | 0.90 | 2.68 |

All variables significantly associated with pathological IMT or Plaques at the age- and sex-adjusted analysis were included in the model.
**Bold:** p<0.05

## Discussion

As previously seen, HIV-infected individuals appear to be at higher risk of CVD than the general population [1–2]. Particularly, chronic inflammatory processes are activated, and atherosclerosis is accelerated [7, 10, 11] in HIV patients; this is the reason why cardiovascular disease is one of the most common non-AIDS events with overall increased morbidity and mortality. Although the mechanisms involved remain elusive, endothelial activation due to the chronic inflammation seems to be the keystone of this phenomenon; in which proinflammatory cytokines [7], pro-angiogenic hematopoietic and endothelial progenitor cells [10], circulating CD40 ligand, and Dickkopf-1 [11] could be involved.

Carotid IMT and presence of plaque have been shown to predict cardiovascular events in large studies [12, 13]. Also, in asymptomatic patients without CVD, carotid IMT and plaque assessment are more likely to revise Framingham Risk Score than Coronary Artery Calcium score. [14]. Furthermore, common carotid blood flow (CBF) velocity was independently associated with future cardiovascular disease (CVD) using color duplex ultrasound and Doppler spectral analysis [15]. In clinical practice, evaluation of the carotid artery by ultrasonography is a very useful, simple, and safe method to indirectly detect and prevent CVD. In preventive medicine, IMT measurement is especially important for subjects with an intermediate CV risk, i.e., for subjects with a 10-year risk of CV disease between 6% and 20%. [16].

Our data shows that the overall percentage of PLWH with carotid impairments, either IMT or plaques, nowadays remains high (30.2%). In fact, in spite of the great improvement of antiretroviral therapies, including the general amelioration of the quality of life among PLWH, and the increase of their life expectancy, this percentage does not seem to show substantial improvement in the last decades compared with our previous observations. As a matter of fact, a percentage of carotid lesions of 35.2% was observed in 2000, [4] 31.7% in 2004 [5], while, in 2017, in a prospective study on advanced naïve subjects, for a population at higher risk of CV disease, the lesions were observed in 38.7% of the patients, after one year of antiretroviral therapy [7]. In all these studies, patients treated with protease inhibitors showed a significantly higher percentage of lesions, and this was in line with data deriving from D:A:D cohort [17]. It can be hypothesized that if, in the past, the increased CV risk of PLHW was due to the HIV-related inflammation, nowadays the vascular damage is sustained by the increase of the average age, and by the higher incidence of age-related comorbidities, despite the advent of new antiretrovirals, more effective in controlling the infection and the consequent inflammation. In our experience, several classic risk factors (older age, male sex, overweight, hypertension, high systolic blood pressure, high blood glucose, diabetes, previous myocardial infarction, low HDL-cholesterol, and high triglycerides) seems to play a central role in carotid impairments. Not unexpectedly, patients with liver and kidney disease seem to be at increased risk of carotid impairments. Variables related to HIV infection (HIV infection duration and CD4+ cell nadir) still exerted a role; in our previous experience, CDC classification [4] and CD4+ cell count [5] were significantly related to epi-aortic impairments. In the present study, the outcome of naïve condition was protective, although the number of such patients was very low.

By analyzing separately IMT and plaques, it was shown that both were significantly related to male sex and age, while HCV infection was associated only with IMT. BMI, hypertension, diabetes, HDL-c, triglycerides, CD4+ cell nadir were related only to plaques. As expected, plaques provided us more information regarding the determinants of vascular damage, with respect to IMT that remains, nonetheless, a sensitive marker of an initial endothelial injury.

Considering the role of antiretroviral therapies, differently from our previous papers where the role of PI constantly emerged [4, 5, 7], our present study evidenced that carotid lesions were related only to current use of raltegravir. Also, by analyzing separately IMT and plaques, the role of the current use of raltegravir emerged for both. This is consistent with recent evidences showing that integrase inhibitors can determine weight gain in PLWH [18, 19]; moreover, a study in 2018 [20] described increase in waist circumference in patients treated with raltegravir use. On the other hand, a correlation between short-term weight gain, subsequent risk of cardiovascular disease, and diabetes, has been evidenced in the D:A:D: cohort [21]. The reason why we observed this correlation only for raltegravir may lie in the fact that the sample size of patients treated with elvitegravir or dolutegravir in our cohort is smaller, and dolutegravir approval is more recent than raltegravir. It was also observed that the past use of Non-Nucleoside Reverse Transcriptase Inhibitors was associated with a lower risk of plaques. This seems homogenous with a body of evidence that confirmed over time the cardiovascular safety of this class of antiretrovirals [22–24].

The main strengths of the present study are the large number of patients involved, and the fact that the cohort involves centers from all the national territory. The major limitation is that, at present, only cross-sectional data are available.

In conclusion, in our experience, among PLWH, color-Doppler ultrasonography could play a pivotal role in identifying and quantifying atherosclerotic lesions, even at a very premature stage, and should be included in the algorithms of comorbidity management of these patients. Our data suggests that a registry of echoic images deriving from all the national territory represents an important source of data, which will allow us to be able to track the CV risk of PLWH and to evaluate the modifications over the time of the role of the different risk factors, both traditional and related to HIV infection, and also the impact of the new and old antiretroviral regimens on CV risk.

## Data Availability

All data produced in the present study are available upon reasonable request to the author

## Acknowledgements

PM designed the study, analyzed, interpreted the data and wrote the manuscript;

AC and YS analyzed and interpreted the data;

CM and LG acquired the data and reviewed the manuscript

EDR performed the statistical analysis;

BMC, SF, RB, GDF, FT and AC collected the data from the participant centers.

